# Predictors of Anxiety Regarding The COVID-19 Pandemic Among Health-care Workers in a Hospital Not Assigned to Manage COVID-19 Patients in Nepal

**DOI:** 10.1101/2020.07.08.20148866

**Authors:** Lekhjung Thapa, Aparna Ghimire, Sulochana Ghimire, Nooma Sharma, Shakti Shrestha, Medha Devkota, Suman Bhattarai, Anzil Man Singh Maharjan, Subash Lohani, Subash Phuyal, Pratibha Maharjan

## Abstract

**Introduction:** We studied the levels and severity of anxiety caused by COVID-19 amongst frontline health-care workers (HCWs) in a tertiary care neurological hospital in Nepal, not assigned to manage COVID-19 cases.

**Methods:** A cross-sectional study was conducted on 100 frontline Health Care Workers (HCWs) using a 10-point subjective assessment tool, the Anxiety Level Index (ALI), and the Zung Self Rating Anxiety Score (SAS), to assess the level of anxiety regarding COVID-19.

**Results:** On ALI 55% of HCW were found to have marked severe anxiety (6-9), however, on SAS 44% of HCW’s reported anxiety. The majority HCW’s were female (n=78) with nurses forming 62% of the sample size. The mean age (±SD) was 26.8 years (± 8.17). Factors associated with significantly higher levels of anxiety regarding COVID-19 on ALI were age (p=0.002), sex (p=0.001), receiving regular COVID-19 updates via social media (p=0.011) and a high frequency of checking for COVID-19 information authenticity (p=0.039). Work experience (p=0.026) and frequency of checking for information authenticity (p=0.029) were factors found to increase SAS measured anxiety and were found to be associated with significantly higher levels of anxiety. Multivariate logistic regression analysis showed that respondents with work experience of ≤2 years were 0.380 (95% CI 0.158 to 0.910) times less likely to have anxiety than those with work experience of ≥2 years. Similarly, the odds of having anxiety among those who checked information authenticity less frequently was 0.377 (95% CI 0.153 to 0.931) times less than those who often did.

**Conclusion:** The COVID-19 outbreak has caused a substantial impact on the mental health of frontline HCWs in a hospital not assigned to manage COVID-19 patients. Length of time of worked in healthcare and the frequency of checking for COVID-19-related information were significant predictors of anxiety.

## Introduction

The novel coronavirus (2019-nCov) was identified in late 2019 in Wuhan city, Hubei province of China. It primarily causes mild to severe respiratory problems, the disease associated with it is termed as COVID-19.^1^ Since it was detected, it has infected over 2 million people and taken more than 120 thousand lives globally.^2^ At the time of data collection (March 23 and 24), the pandemic had resulted in 51,862 cases and 1,941 deaths globally.^3^ The number of active cases in Nepal was only one (01).^4^ However when this manuscript was being prepared Nepal had reported total 52 cases, among whom 16 had recovered leaving 36 active cases as of April 26, 2020.^5^

The scarcity of information about this novel virus, lack of medical evidence regarding the treatment and prevention of the disease, the exponential rise in the number of infected people, and the increasing death toll have created global alarm and anxiety.^6,7^ Figures from China’s National Health Commission reveal that more than 3300 HCWs were infected by early March and according to local media, by the end of February at least 22 had died. In Italy, 20% of responding HCWs were infected, and some had died.^8^ Reports about infection among HCWs from the United States are also alarming, with 9,282 HCWs infected by April, 2020.^9^ These facts have resulted in high levels of anxiety, especially among frontline HCW all over the world.

In a recently published report by Lai et al., frontline HCWs engaged in the management of patients with COVID-19 in high risk environments were found to have a significantly higher risk of developing anxiety.^10^ In a report from Wuhan during the peak of the outbreak, frontline HCWs were under moderate to severe stress, and many had elevated levels of anxiety and depression.^11^

Similar phenomenon was observed during the Severe Acute Respiratory Syndrome (SARS) pandemic, which caused significant psychological stress among HCWs.^12^ Interestingly the level of perceived psychological stress was found to be similar among HCWs working in high-risk environments, compared to those working at lower risk environments^13^. Similar findings can be expected in frontline HCWs not engaged in direct patient contact, diagnosis and treatment with COVID-19, primarily as during the pandemic all patients admitted to a hospital are suspected of having COVID-19. Therefore, there is a high probability that frontline HCWs are more likely to expose themselves to COVID-19 in hospital.

Importantly, out of 4,282 registered public health facilities^14^ only 25 hospitals in Nepal have been assigned to manage COVID-19 cases by the Government. Therefore, there are a high proportion of frontline HCWs who are not engaged in the direct care of COVID-19 patients in Nepal. Majority of these HCWs are likely to be struggling with debilitating stress and anxiety despite not being in clinical contact with COVID-19 patients. As a result, they may be underperforming in their routine job of caring for patients who have tested –ve for COVID-19. Therefore, in this group of HCWs, assessment of psychological distress in the form of anxiety assessments would be an essential first step to understand their current mental status. This could help guide us to plan the intervention, if required.

We therefore, have studied COVID-19 related anxiety and its severity in frontline HCWs of Upendra Devkota Memorial-National Institute of Neurological and Allied Sciences (UDM-NINAS), Kathmandu, which is currently not designated for the care of patients with COVID-19 in Nepal.

## Methods

### Design

In this cross-sectional study, we assessed the level of anxiety related to the COVID-19 pandemic among frontline health-care workers (HCWs) at UDM-NINAS, using a 10-point Anxiety Level Index (ALI) and the Zung Self Rating Anxiety Scale (SAS).

### Sample and setting

UDM-NINAS is a 100-bed tertiary care neurological hospital located in the capital city of Nepal, Kathmandu. The hospital has 265 staff, 250 of whom are engaged in direct patient care. We invited all the HCWs to participate in this study, the inclusion criteria were if they were frontline HCWs, and able to read and write English. We defined frontline HCWs as those working as doctors, nurses, physiotherapists, lab technicians, radiology technicians, administrative staff, and pharmacists. We enrolled a total of 100 participants. Those not willing to participate were excluded from the study.

### Measurements

#### Sociodemographic data and questionnaire related to COVID-19 information

Before the assessment, participants provided data on their age, sex, education level, permanent address, job title, experience in the medical field, and experience at UDM-NINAS. They also responded to a questionnaire consisting of five questions related to COVID-19.

#### Anxiety

For this study, anxiety was defined as a transient emotional state consisting of feelings of apprehension, nervousness, and physiological sequelae such as an increased heart rate or respiratory rate.^16^

Each respondent completed two self-reported questionnaires that reflected the state anxiety: (1) Anxiety Level Index (ALI); and (2) Zung Self-Rated Anxiety Scale (SAS). The ALI is an investigator designed one-item, numeric rating instrument. The participant reads the following statement: “On a scale of 0-10 (0 being no anxiety at all and 10 being the most anxiety you have ever experienced), how much do you rate your anxiety related to COVID-19 currently?” The responses were then categorized as no anxiety (Score: 0); minimal to moderate (Score: 1-5); marked to severe (Score: 6-9); and most extreme anxiety (Score: 10). The responses were further categorized into two groups: “Group 1” as having “no anxiety” (Score: 0); and “Group II” as having “anxiety” (Score: 1-10).

The SAS is a 20-item instrument that enables participants to rate their current levels of anxiety, covering both psychological and somatic symptoms. For each item, respondents indicated their response using a scale of 1 (None or a little of the time) to 4 (Most or all the time). Thus, the total raw scores ranged from 20 to 80. The instrument is less time consuming (5 to 10 minutes to complete) and has been used to assess anxiety in previous studies.^17^ For our study sample, Cronbach’s α reliability coefficient was 0.79. The raw score obtained was converted to Anxiety Index (AI) using the conversion tool.^18^ Finally, based on the AI, the participants were categorized as: no anxiety (AI: 20-44); minimal to moderate (AI: 45-59); marked to severe (AI; 60-74); and most extreme anxiety (AI ≥75).^19^ We further categorized the respondents into two groups as: “Group 1” having “no anxiety” (AI: 20-44); and “Group II” as having “anxiety” (AI ≥ 45).

### Procedure

The Institutional Review Board approved the study. Before data collection, all participants gave informed, written consent. The investigator (AG) approached all the staff who met the inclusion criteria to fill their responses in a proforma. Data was collected within two hours of completion of the proforma.

### Statistical analysis

Participant sociodemographic information, responses to COVID-19 questionnaire, and anxiety were characterized using descriptive statistics. While all variables were converted to dichotomous values prior to bivariate and multivariate analysis, the severity of anxiety was not only confined to bivariate analysis. Bivariate analysis was used to determine the association of the outcome variable (anxiety) with an individual explanatory variable (sociodemographic variables and response to COVID-19 questions). Multivariate binary logistic regression analysis using backward stepwise likelihood ratio method was used to determine the influence of all the explanatory variables (significant on the bivariate analysis) on anxiety (dichotomous). The statistically significant value was set at <0.05 for all the analyses.

## Results

Out of 100 respondents, the majority were female (n=78), with majority being nurses (62%). The mean age (±SD) of the respondents was 26.8 years (± 8.17), while 79% were in the age group of 20-29 years. 38% were permanent residents of Kathmandu valley and 51% were undergraduates. Slightly over 3/4^th^ respondents had ≤5 years of experience in the medical field (74%), and association with UDM-NINAS (81%). **[Table 1]**

**Table 1:** Socio-demographic variables.

| **Variables** | **Frequency** | **Percentage** |
| --- | --- | --- |
| **Age Category (in Years)** |  |  |
| 0-19 | 1 | 1.0 |
| 20-29 | 79 | 79.0 |
| 30-39 | 13 | 13.0 |
| 40-49 | 3 | 3.0 |
| 50-59 | 2 | 2.0 |
| 60-69 | 2 | 2.0 |
| *Mean Age ± SD (26.8 ± 8.17)* |  |  |
| **Sex** |  |  |
| Female | 78 | 78.0 |
| Male | 22 | 22.0 |
| **Permanent Residence** |  |  |
| Inside Kathmandu Valley | 38 | 38.0 |
| Outside Kathmandu Valley | 62 | 62.0 |
| **Level of Education** |  |  |
| Undergraduate | 51 | 51.0 |
| Graduate | 42 | 42.0 |
| Postgraduate | 7 | 7.0 |
| **Job Specification** |  |  |
| Administrative Staffs | 17 | 17.0 |
| Paramedics | 12 | 12.0 |
| Nurse | 62 | 62.0 |
| Doctor | 9 | 9.0 |
| **Experience in Medical Field (in Years)** |  |  |
| $\leq 5$ | 74 | 74.0 |
| 6-10 | 15 | 15.0 |
| >10 | 11 | 11.0 |
| **Experience in UDM-NINAS (in Years)** |  |  |
| $\leq 5$ | 81 | 81.0 |
| 6-10 | 11 | 11.0 |
| >10 | 08 | 08.0 |

The participants’ responses to questions about information regarding COVID-19 is shown in **Table 2**. On ALI (0-10-point subjective scale), the mean score (±SD) was 6.94 (±2.22), and majority of the participants (55%) reported marked-severe anxiety (6-9). 17% of the participants rated their anxiety as being 10 out of 10. **[Table 3]**

**Table 2:** Participants response regarding information about COVID-19 (n=100)

| **Questions** | **Options** | **Frequency** | **Percentage** |
| --- | --- | --- | --- |
| 1. *When did you first hear about COVID-19?* | < 1month | 26 | 26.0 |
|  | 1-3 months | 74 | 74.0 |
| 2. *How did you first hear about COVID-19?* | Colleague/s | 1 | 1.0 |
|  | Social media | 76 | 76.0 |
|  | News portal | 23 | 23.0 |
| 3. *How frequently do you update yourself about COVID-19?* | <1 time/day | 7 | 7.0 |
|  | 1-5 times/day | 51 | 51.0 |
|  | >5 times/day | 20 | 20.0 |
|  | >every hour | 22 | 22.0 |
| 4. *Which medium do you use for information about COVID-19? (multiple answers)* | Social media | 77 | 77.0 |
|  | News portal | 28 | 28.0 |
|  | WHO website | 36 | 36.0 |
|  | Worldometer | 8 | 8.0 |
|  | Medical journal | 6 | 6.0 |
| 5. *How frequently do you check the authenticity of the information?* | Never | 2 | 2.0 |
|  | Sometimes | 44 | 44.0 |
|  | Most of the time | 47 | 47.0 |
|  | Always | 7 | 7.0 |

**Table 3:**
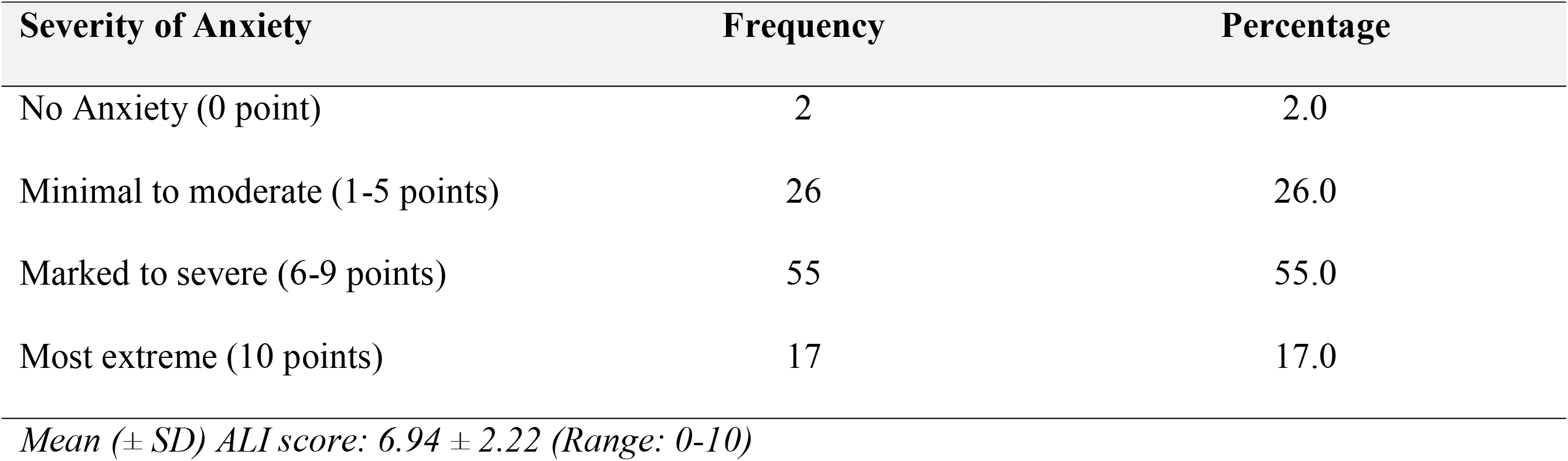
Severity of Anxiety detected by ALI (0-10-point score) (n=100)

| Severity of Anxiety | Frequency | Percentage |
| --- | --- | --- |
| No Anxiety (0 point) | 2 | 2.0 |
| Minimal to moderate (1-5 points) | 26 | 26.0 |
| Marked to severe (6-9 points) | 55 | 55.0 |
| Most extreme (10 points) | 17 | 17.0 |

*Mean ( ± SD) ALI score: $6.94 \pm 2.22$ (Range: 0-10)*

On the SAS, 66% of respondents had normal scores (20-44) implying no anxiety and 28% had mild to moderate levels of anxiety (45-59). Six had marked to severe levels of anxiety. **[Table 4]** Table 5 and 6 show the differences in sociodemographic variables and responses to questions on COVID-19 according to the severity of anxiety (ordinal) measured by ALI and SAS, respectively. **Table 5** suggests that age (p=0.002), sex (p=0.001), being updated on COVID-19 via social media (p=0.011) and frequency of checking for COVID-19 information authenticity (p=0.039) were significantly associated with higher levels of anxiety as assessed by ALI. However, the severity of anxiety assessed by SAS was significantly associated with the permanent residence (p=0.038), job specification (p=0.013), and duration of work experience at UDM-NINAS (p=0.046) **[Table 6]**.

**Table 4:**
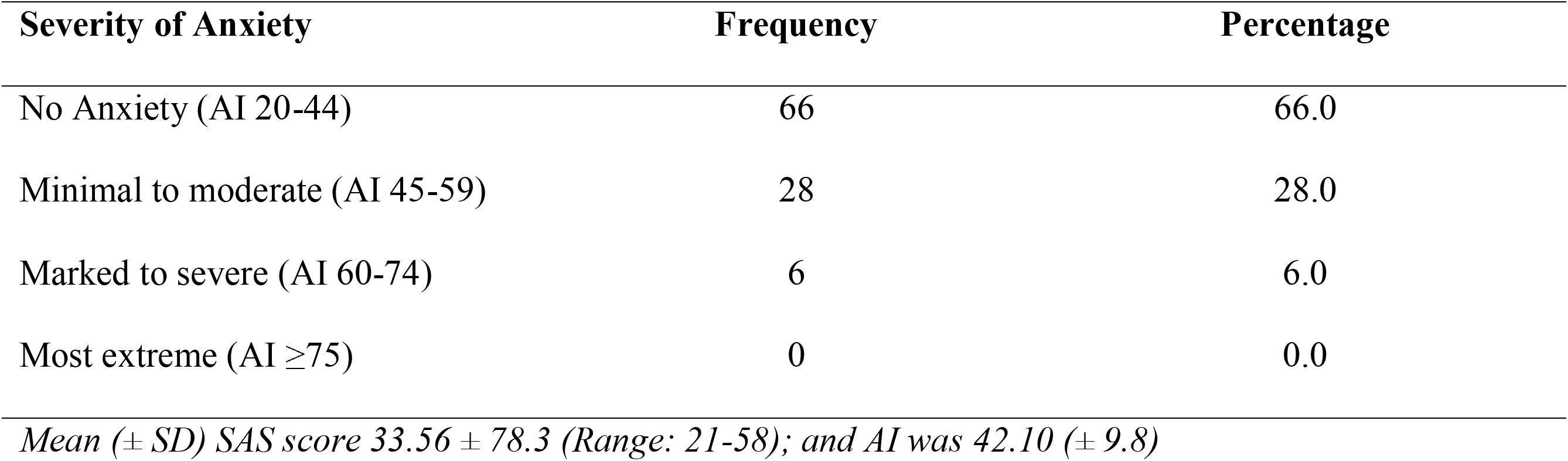
Severity of Anxiety in Zung Self-Rating Anxiety Scale (SAS) (n=100)

| Severity of Anxiety | Frequency | Percentage |
| --- | --- | --- |
| No Anxiety (AI 20-44) | 66 | 66.0 |
| Minimal to moderate (AI 45-59) | 28 | 28.0 |
| Marked to severe (AI 60-74) | 6 | 6.0 |
| Most extreme (AI $\geq 75$) | 0 | 0.0 |

*Mean ( ± SD) SAS score $33.56 \pm 78.3$ (Range: 21-58); and AI was $42.10 (\pm 9.8)$*

**Table 5:**
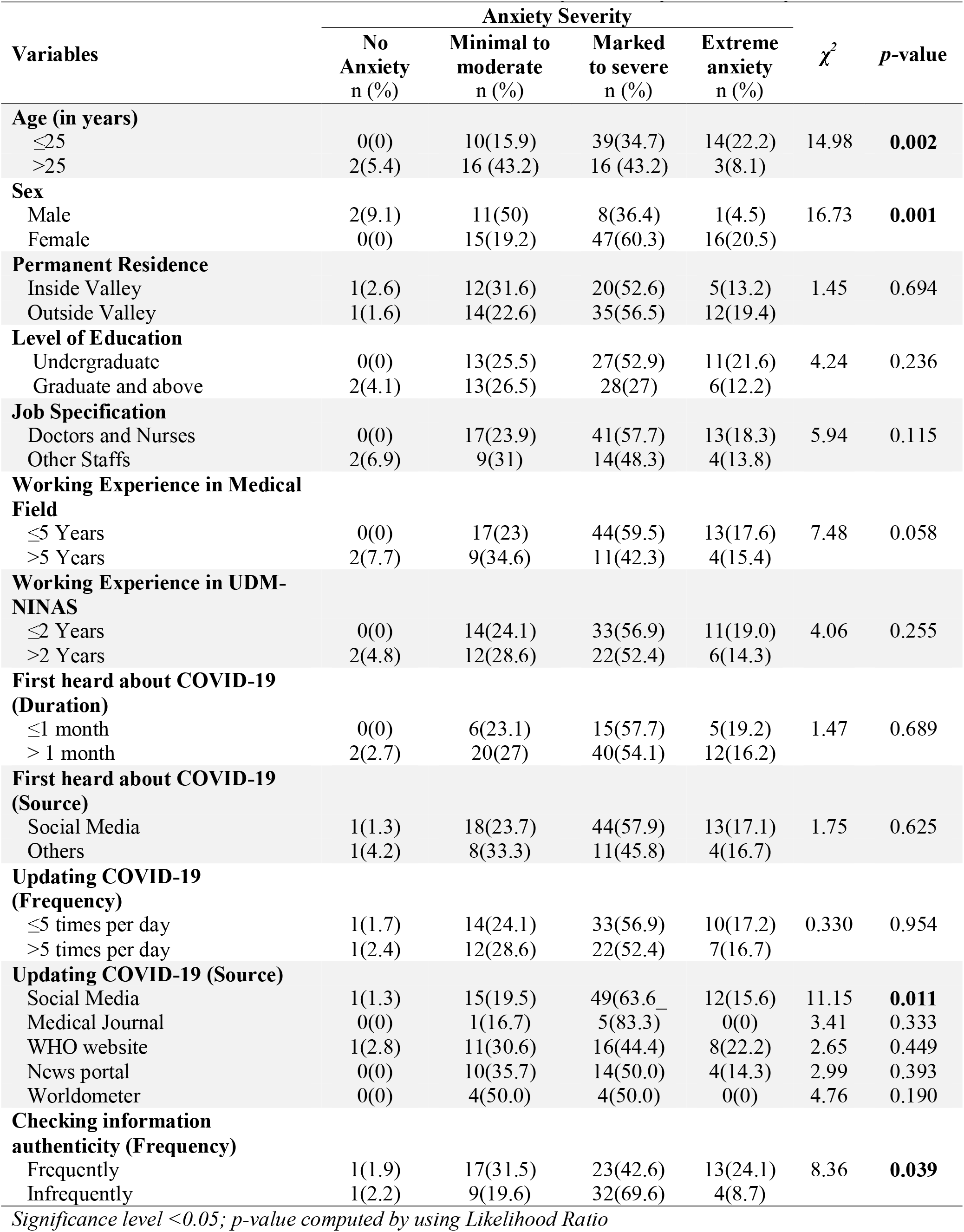
Association Between Baseline Variables and Severity of Anxiety Measured by ALI.

**Table 6:** Association Between Baseline Variables and Severity of Anxiety Measured by SAS.

| Variables | Anxiety Severity | | | | $\chi^2$ | p-value |
| --- | --- | --- | --- | --- | --- | --- |
|  | No Anxiety<br>n (%) | Minimal to moderate<br>n (%) | Marked to severe<br>n (%) | Extreme anxiety<br>n (%) |  |  |
| **Age (in years)** |  |  |  |  |  |  |
| ≤25 | 41(65.1) | 18(28.6) | 4(6.3) | 0(0) | 0.08 | 0.962 |
| >25 | 25(67.6) | 10(27.0) | 2(5.4) | 0(0) |  |  |
| **Sex** |  |  |  |  |  |  |
| Male | 16(72.7) | 6(27.3) | 0(0) | 0(0) | 3.18 | 0.204 |
| Female | 50(64.1) | 22(28.2) | 6(7.7) | 0(0) |  |  |
| **Permanent Residence** |  |  |  |  |  |  |
| Inside Valley | 25(65.8) | 13(34.2) | 0(0) | 0(0) | 6.56 | **0.038** |
| Outside Valley | 41(66.1) | 15(24.2) | 6(9.7) | 0(0) |  |  |
| **Level of Education** |  |  |  |  |  |  |
| Undergraduate | 34(66.7) | 13(25.5) | 4(7.8) | 0(0) | 0.84 | 0.656 |
| Graduate and above | 32(65.3) | 15(30.6) | 2(4.1) | 0(0) |  |  |
| **Job Specification** |  |  |  |  |  |  |
| Doctors and Nurses | 50(70.4) | 15(21.1) | 6(8.5) | 0(0) | 8.65 | **0.013** |
| Other Staffs | 16(55.2) | 13(44.8) | 0(0) | 0(0) |  |  |
| **Working Experience in Medical Field** |  |  |  |  |  |  |
| ≤5 Years | 53(71.6) | 18(24.3) | 3(4.1) | 0(0) | 4.30 | 0.116 |
| >5 Years | 13(50) | 10(38.5) | 3(11.5) | 0(0) |  |  |
| **Working Experience in UDM-NINAS** |  |  |  |  |  |  |
| ≤2 Years | 44(75.9) | 12(20.7) | 2(3.4) | 0(0) | 6.16 | **0.046** |
| >2 Years | 22(52.4) | 16(38.1) | 4(9.5) | 0(0) |  |  |
| **First heard about COVID-19 (Duration)** |  |  |  |  |  |  |
| ≤1 month | 16(61.5) | 7(26.9) | 3(11.5) | 0(0) | 1.69 | 0.429 |
| > 1 month | 50(67.6) | 21(28.4) | 3(4.1) | 0(0) |  |  |
| **First heard about COVID-19 (Source)** |  |  |  |  |  |  |
| Social Media | 49(64.5) | 21(27.6) | 6(7.9) | 0(0) | 3.42 | 0.181 |
| Others | 17(70.8) | 7(29.2) | 0(0) | 0(0) |  |  |
| **Updating COVID-19 (Frequency)** |  |  |  |  |  |  |
| ≤5 times per day | 36(62.1) | 17(29.3) | 5(8.6) | 0(0) | 2.18 | 0.336 |
| >5 times per day | 30(71.4) | 11(26.2) | 1(2.4) | 0(0) |  |  |
| **Updating COVID-19 (Source)** |  |  |  |  |  |  |
| Social Media | 51(66.2) | 20(26.0) | 6(7.8) | 0(0) | 3.61 | 0.165 |
| Medical Journal | 5(83.3) | 0(0) | 1(16.7) | 0(0) | 4.57 | 0.102 |
| WHO website | 26(72.2) | 8(22.2) | 2(5.6) | 0(0) | 4.04 | 0.595 |
| News portal | 21(75.0) | 4(14.3) | 3(10.7) | 0(0) | 4.74 | 0.093 |
| Worldometer | 6(75.0) | 1(12.5) | 1(12.5) | 0(0) | 1.51 | 0.471 |
| **Checking information authenticity (Frequency)** |  |  |  |  |  |  |
| Frequently | 30(55.6) | 20(37) | 4(7.4) | 0(0) | 5.90 | 0.052 |
| Infrequently | 36(78.3) | 8(17.4) | 2(4.3) | 0(0) |  |  |

Significance level <0.05; p-value computed by using Likelihood Ratio

An explorative analysis of potential factors influencing anxiety (dichotomous) measured by both ALI and SAS were attempted, but the multivariate logistic regression model for the former was invariably distorted with high standard error; consequently, only a bivariate analysis has been presented for ALI measured anxiety **[Table 7]** whereas both have been presented for SAS related anxiety **[Table 8]. Table 7** shows that there was a statistically significant association between ALI assessed anxiety and sex (p=0.047) on bivariate analysis. The explorative analysis of factors influencing SAS measured anxiety among frontline HCWs at UDM-NINAS upon a bivariate analysis suggests that the experience of working in a hospital (p=0.026) and checking for information authenticity (p=0.029) were significantly but independently associated with higher levels of anxiety. However, the multivariate logistic regression analysis showed that those who had experience of working in the hospital for two years or less were 0.380 (95% CI 0.158 to 0.910) times less likely to have anxiety than those working for two years and more.

**Table 7:** Association Between Baseline Variables and Anxiety Measured by ALI.

| Variables | Anxiety | | $\chi^2$ | p-value |
| --- | --- | --- | --- | --- |
|  | Absent<br>n (%) | Present<br>n (%) |  |  |
| **Age (in years)** |  |  |  |  |
| ≤25 | 0(0) | 63(100) | 1.26 | 0.135 |
| >25 | 2(5.4) | 35(94.6) |  |  |
| **Sex** |  |  |  |  |
| Male | 2 (9.1) | 20(90.9) | 3.34 | **0.047\*** |
| Female | 0(0) | 78(100) |  |  |
| **Permanent Residence** |  |  |  |  |
| Inside Valley | 1(2.6) | 37(97.4) | <0.01 | 1.000 |
| Outside Valley | 1(1.6) | 61(98.4) |  |  |
| **Level of Education** |  |  |  |  |
| Undergraduate | 0(0) | 51(100) | 0.55 | 0.238 |
| Graduate and above | 2(4.1) | 47(95.9) |  |  |
| **Job Specification** |  |  |  |  |
| Doctors and Nurses | 0 (0) | 71(100) | 2.10 | 0.082 |
| Other Staffs | 2(6.9) | 27(93.1) |  |  |
| **Working Experience in Medical Field** |  |  |  |  |
| ≤5 Years | 0(0) | 74(100) | 2.55 | 0.066 |
| >5 Years | 2(7.7) | 24(92.3) |  |  |
| **Working Experience in UDM-NINAS** |  |  |  |  |
| ≤2 Years | 0(0) | 58(100) | 0.91 | 0.174 |
| >2 Years | 2(4.8) | 40(95.2) |  |  |
| **First heard about COVID-19 (Duration)** |  |  |  |  |
| ≤1 month | 0(0) | 26(100) | <0.01 | 1.000 |
| > 1 month | 2(2.7) | 72(97.3) |  |  |
| **First heard about COVID-19 (Source)** |  |  |  |  |
| Social Media | 1(1.3) | 75(98.7) | <0.01 | 0.424 |
| Others | 1(4.2) | 23(95.8) |  |  |
| **Updating COVID-19 (Frequency)** |  |  |  |  |
| ≤5 times per day | 1(1.7) | 57(98.3) | <0.01 | 1.000 |
| >5 times per day | 1(2.4) | 41(97.6) |  |  |
| **Updating COVID-19 (Source)** |  |  |  |  |
| Social Media | 1(1.3) | 76(98.7) | <0.01 | 0.409 |
| Medical Journal | 0(0) | 6(100.0) | <0.01 | 1.000 |
| WHO website | 1(2.8) | 35(97.2) | <0.01 | 1.000 |
| News portal | 0(0) | 28(100.0) | 0.01 | 1.000 |
| Worldometer | 0(0) | 8(100.0) | <0.01 | 1.000 |
| **Checking information authenticity (Frequency)** |  |  |  |  |
| Frequently | 1(1.9) | 53(98.1) | <0.01 | 1.000 |
| Infrequently | 1(2.2) | 45(97.8) |  |  |

Significance level <0.05; p-value computed by using Fischer's Exact Chi-square test

**Table 8:**
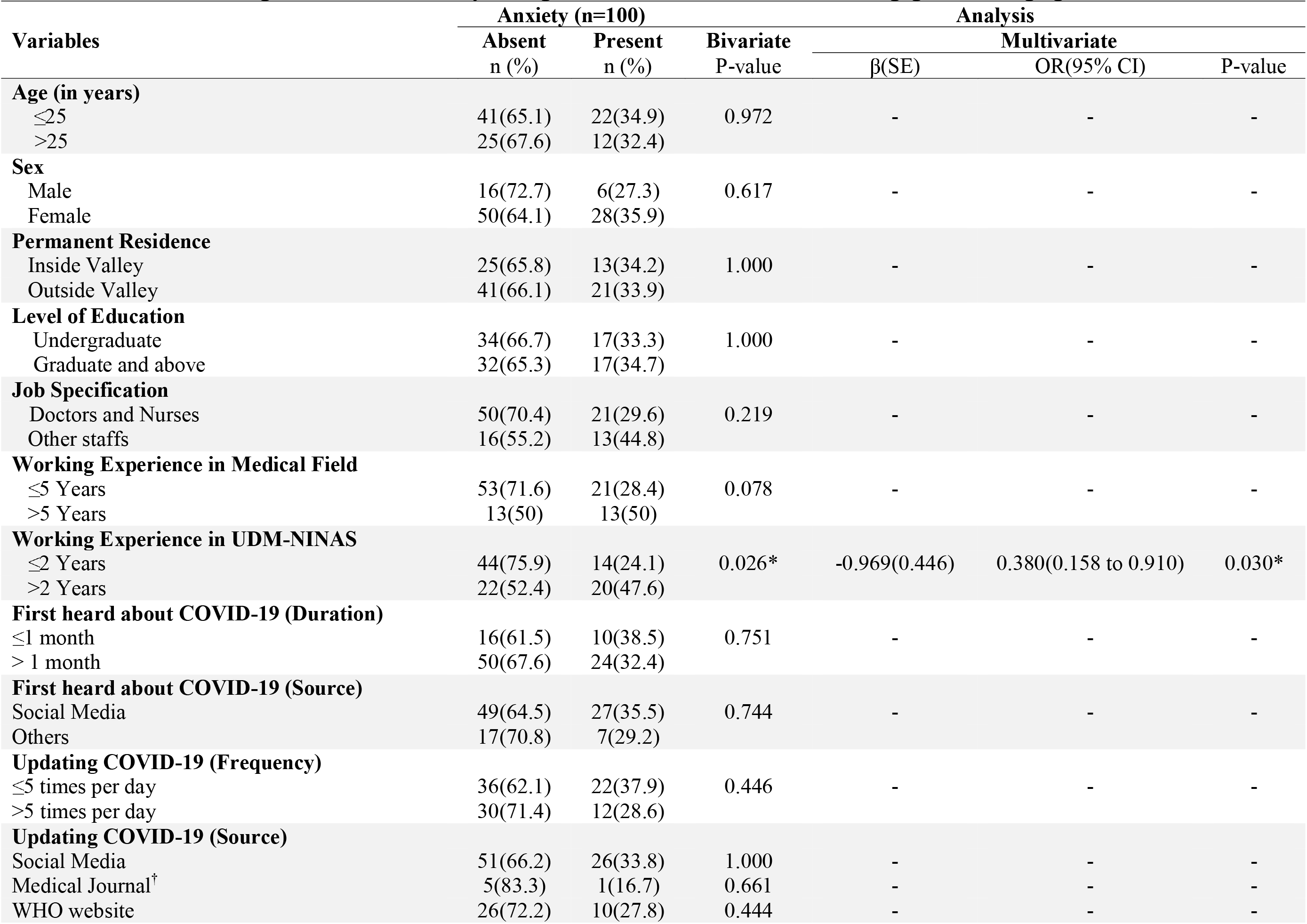
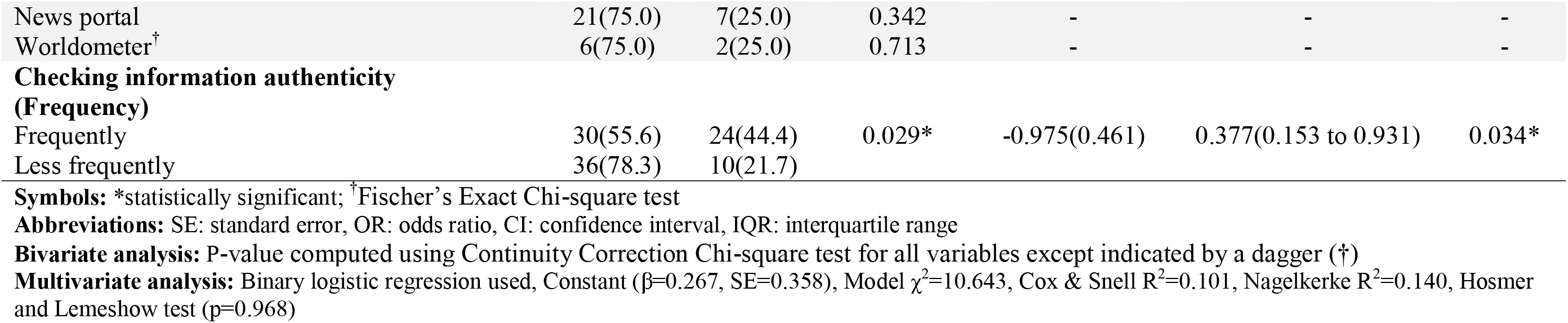
Factors influencing SAS measured anxiety among frontline health-care workers not engaged in managing COVID-19 cases.

Similarly, the odds of having anxiety among those who less often checked for information authenticity was 0.377 (95% CI 0.153 to 0.931) times less than those who frequently checked for COVID-19 related information authenticity.

## Discussion

This study showed that a high proportion of respondents felt anxious due to the COVID-19 pandemic. Even during routine work, professionals in the healthcare industry have relatively higher levels of stress compared to other professions^20^ resulting in higher rates of chronic stress, depression and anxiety.^12, 21, 22^ Research from Singapore reported anxiety to be prevalent in 23% of all HCWs.^23^ In 2003, during the SARS pandemic, a situation similar to the COVID-19 pandemic, Poon et al. from Hongkong reported higher anxiety scores amongst doctors and administration staff.^24^ Notably, in the same study, anxiety levels in HCWs not in direct contact with SARS patients were also found to be significantly high,^24^ as observed among the respondents of our study who were not in direct contact with COVID-19 cases.

### COVID-19-related information among the respondents

In this study, approximately 1/4^th^ respondents had heard about COVID-19 for less than a month. COVID-19 related pneumonia was diagnosed and reported to the WHO on 31^st^ December 2019, three months before our data collection. Although the WHO was concerned about the disease and declared the outbreak a public health emergency of international concern on 30^th^ January 2020,^25^ it was only on 11^th^ March 2020 that the WHO declared it a pandemic.^26^ This delay in declaration by the concerned authorities may be one of the reasons why only a small proportion of our respondents knew about the disease for less than a month. Majority of respondents (76%) had first heard about COVID-19 on social media and an almost similar proportion of the respondents used social media to find out more information about COVID-19.

There are now 3.81 billion social media users worldwide, representing 49% of the world’s total population.^27^ Social media in the current era has become an important tool for governments, non-governmental organisations, and individuals to exchange valuable information.^28^ It was observed that during this pandemic messaging on Instagram and Facebook soared by over 50 percent in many countries, people were found to repeatedly click on virus related news stories on social media.^29^

Slightly over half (51%) of the respondents reported checking for new COVID-19-related information 1-5 times a day. Moreover, curiosity and anxiety about the pandemic increased exponentially due to implementation of a nationwide lockdown in Nepal, only one day before the commencement of our data collection. Purgato et al. have reported the use of social media for the critical updates as being associated with higher levels of acute stress during University lockdown.^30^

Majority of the participants in our study used social media for information updates and only about 1/3^rd^ (36%) of our respondents visited the WHO website for information. This finding suggests that people preferred using social media for updates regarding COVID-19 since most social media platforms are easy to use, requiring no technical expertise and mobile phones with internet access are now widely available making this information easy to access.

Globally, COVID-19 has been an unprecedented public health crisis and the use of social media platforms like Facebook, Twitter, YouTube, Instagram etc. are at the heart of this crisis facilitating easy access and distribution of Covid-19 related information. However, this pandemic has also shown how easy it is for false information to spread through these platforms. Even the WHO noted that urgent measures needed to be taken to address “corona infodemics.”^31^ In a study exploring sharing information on social platforms, Krishna R. found that only 6% respondent verified the information before sharing in social media.^32^

Therefore it seems prudent to check the authenticity of the information obtained from social media. Over half of the respondents (54%) reported to always, or in the majority of times check the authenticity of the information.

### ALI, SAS and anxiety

COVID-19 pandemic related anxiety has the potential to prevent HCWs from caring for patients in the best possible manner due to increased amounts of work-related stress. Nearly all of our respondents (98%) had anxiety as assessed by ALI, and 17% were found to have extreme levels of anxiety. However, SAS revealed that only 34% of our respondents had anxiety related symptoms. This discrepancy could be because ALI captures the feeling of anxiety in a visual analogue scale, however SAS assesses the symptoms related to anxiety in a comprehensive manner. Different comprehensive anxiety scales are have been used to assess anxiety, the Likert scale to assess anxiety has been suggested as an adequate replacement for tools like the State Trait Anxiety Inventory (STAI) to assess for current anxiety.^33^ However to interpret if ALI can be used as an alternative to SAS to assess anxiety is beyond the scope of this study.

A Chinese study during the COVID-19 pandemic reported a slightly higher number of HCWs (44.6%) to have anxiety symptoms,^12^ as compared to our study (34%). A study from Beijing during the SARS outbreak reported 10% of hospital employees as having high SARS related post-traumatic stress (PTS) score.^34^ This discrepancy can be due to research in China being carried out in hospitals where COVID-19 patients were undergoing treatment and the large number of infected people in the country. However, in our case the data was collected when there were only two positive cases of COVID-19 in the country and both had been imported from abroad.^35^ Additionally the tools we used to assess anxiety were different.

In the study from China being female, nurse, and intermediate technical title were identified to be significant predictors of the severity of symptoms.^12^ Similar to these findings, our study found that females with self-reported anxiety were found to have increased severity of symptoms as compared to males in both ALI and SAS and majority of the females were nurses. The female gender was significantly associated with higher levels of anxiety as assessed by ALI.

Across the world physicians, nurses, and other frontline HCWs have put their lives at risk to treat patients with COVID-19 in stressful settings daily.^36,37^ In this study, all the doctors and nurses reported anxiety on ALI. However, only about 1/3^rd^ (29.6%) doctors and nurses, had anxiety as measured by SAS. The reasons for this difference are likely because of the distinctive characteristics of these two scores, as discussed earlier. Shorter duration of work experience was significantly associated with increased anxiety as measured by SAS. A higher proportion of respondents aged ≤25 years old (22.2%) as compared to >25years old (8.1%) had extreme anxiety and younger age was significantly associated with the severity of anxiety on ALI. Younger age and people in the early stages of their careers as HCWs were found to have increased levels of COVID-19 related stress. Maturity is therefore considered to be a protecting factor against stress.^38,39^

A higher proportion of females (20.5%) as compared to males (4.5%) were found to have extreme anxiety, and gender was significantly associated with the severity of symptoms of anxiety on ALI. Research has revealed that female respondents have more negative alterations in cognition or mood sub-symptoms compared to males.^40^

None of the respondents had extreme anxiety on SAS. However permanent residence, job specification, and work experience at the current institute were significantly associated with severity of anxiety measured by SAS. Studies have revealed that years of employment, age, sex, socio-economic status, and individual characteristics such as personality, subjective experiences, emotional maturity have been established as causal factors for psychiatric morbidity.^38,39,40^

Multivariable logistic regression analysis revealed that duration of employment and frequency of checking for authentic of COVID-19-related information were significant predictors of anxiety. Respondents with short duration of employment had higher odds of having anxiety. Koinis A et al. in their study about coping mechanisms among health-care workers observed that professionals who had been employed for longer durations more often had developed positive approach strategies such as problem solving to better deal with work related stress.^42^

Respondents who checked for authenticity of COVID-related information less frequently had lesser odds of having anxiety as compared to those who frequently checked for authenticity of information. This finding is though provoking and complex, as news media exposure alone is not necessarily associated with higher levels of anxiety. However, people who are stressed by an influx of new information and are more likely to check for authenticity of information frequently, as seen in our study are more likely to develop anxiety. We believe that there are several factors influencing this such as the individual’s personal beliefs and level of education which may influence their interpretation of the news. Based on the findings of a study exploring news media and psychological distress, it has been explained that “news media exposure might even provide information which diminishes anxiety”.^43^ There is also the possibility that HCWs who less frequently checked for authenticity of information may have individual characteristics such as increased resilience and more efficient coping strategies both of which are positive psychological traits which lead to positive outcomes in education and mental health.^44^

### Limitations and future directions

Although our study gives information about the anxiety regarding COVID-19 among HCWs not engaged in managing COVID-19 cases in Nepal, it has a few limitations. We intended to enroll all the HCWs in UDM-NINAS; however, because of the lockdown we only could collect data from 100 HCWs, which represents over 50% of the total HCWs of the hospital. Anxiety is a complex phenomenon^45^ and incorporation of essential variables like marital status, partner support, relationship with employer and colleagues, cultural background etc., which can heavily influence anxiety, is suggested. Future longitudinal studies are needed to understand chronic courses and prognosis of anxiety following COVID-19 in these individuals.

## Conclusion

To conclude, the outbreak of COVID-19 has had a significant impact on the mental health of frontline HCW’s in a tertiary neurological center, not assigned to manage COVID-19 infected patients, by increasing the subjective feeling of anxiety as well as expression of anxiety symptoms as assessed by ALI and SAS. Work experience and frequency of checking COVID-19 related information authenticity were valuable predictors of anxiety.

## Data Availability

The data would be made available upon appropriate request.

## Data Availability Statement

The datasets used and analyzed for the current study are available from the corresponding author on reasonable request.

## Authors Contribution

LJT, AG, SG and N Sharma were involved in data collection and preparation of manuscript, S Shrestha was involved in analysis and manuscript editing, SB, AM, SP, SP, MD and PM were involved in manuscript preparation, editing and literature review.

## Acknowledgments

We wish to acknowledge the staff of UDM-NINAS for taking part in the study. We thank Dr MD Devkota, Dr Rachana Nakarmi, Ms Pratikshya, Ms Sarah, Mr Kashi and Mr Bidhyakar Adhikari for valuable help from administration of the hospital.

